# A Multicenter Retrospective Study on the Safety and Efficacy of the Surpass Evolve Flow Diverter System for the Treatment of Intracranial Aneurysms

**DOI:** 10.64898/2026.11.06.26367479

**Authors:** Deyuan Zhu, Mingming Zhang, Wenjia Ma, Shu Zhang, Jihu Zhao, Guangsen Cheng, Yibin Fang

**Author notes:** **Corresponding authors**: Yibin Fang. Corresponding, Guangsen Cheng. Corresponding. Deyuan Zhu and Mingming Zhang should be listed as co-first authors. Deyuan zhu. **Contributorship statement**: Conception and design: Deyuan Zhu and Yibin Fang. Data acquisition: Deyuan Zhu, Wenjia Ma, and Shu Zhang. Analysis and interpretation of data: Deyuan Zhu and Jihu Zhao. Critically revising the article: Yibin Fang, Mingming Zhang, and Guangsen Cheng. Yibin Fang, Mingming Zhang, and Guangsen Cheng reviewed the submitted manuscript. Approved the final version of the manuscript on behalf of all authors: Yibin Fang, Mingming Zhang, and Guangsenn Cheng. Statistical analysis: Deyuan Zhu, Wenjia Ma, and Shu Zhang. Administrative/technical/material support: Deyuan Zhu, Jihu Zhao. **Clinical trial number**: This study was registered with ClinicalTrials.gov (Identifier: NCT06731946) on 29 November 2024. **Human Ethics and Consent to Participate declarations**: This study was conducted in accordance with the Declaration of Helsinki and was approved by the Institutional Review Board (IRB) of the coordinating center (Shanghai Fourth People’s Hospital, Tongji University School of Medicine) with approval number 2024235-001. The requirement for informed consent was waived by the IRB due to the retrospective, observational nature of the study and the use of anonymized patient data. **Consent to Participate declaration**: As this was a retrospective study using anonymized clinical data, the requirement for individual patient consent to participate was waived by the institutional ethics review boards. No identifiable patient information is included in this manuscript. **Ethics declaration**: All procedures performed in this study were in accordance with the ethical standards of the institutional research committee and with the 1964 Helsinki Declaration and its later amendments. The study was approved by the IRB of the coordinating center (approval number 2024235-001). **Data availability statement**: All data generated and analyzed during this study are included in this article. The datasets used and/or analyzed during the current study are available from the corresponding author on reasonable request. Due to institutional ethical restrictions (patient privacy and anonymization protocols), data cannot be shared publicly but are accessible for validated research purposes upon reasonable request to the corresponding author (Yibin Fang, M.D.,), subject to approval from the institutional review board.

## Abstract

**Objective:** To evaluate the safety and efficacy of the Surpass Evolve flow diverter system for the treatment of intracranial aneurysms across three medical centers.

**Methods:** A retrospective review was conducted on patients with intracranial aneurysms treated with the Surpass Evolve FDS between March 2023 and May 2024 at three participating centers. Primary outcome measures included the rate of complete aneurysm occlusion (Raymond-Roy class 1) and the incidence of periprocedural major stroke and mortality. Secondary outcomes encompassed follow-up modified Rankin Scale (mRS) scores, imaging assessment results, and complication rates.

**Results:** A total of 132 patients (132 aneurysms) were included. The median age was 60.0 years (IQR 52.5–69.0), and 80 patients (66.6%) were female. The complete occlusion rate was 73.3% (88/120). The rate of major stroke was 2.3%, with no procedure-related mortality. At follow-up, 96.9% of patients had an mRS score of 0–2.

Multivariate analysis identified non-use of an intraprocedural IIb/IIIa inhibitor and the absence of a branch arising from the aneurysm as independent predictors of complete occlusion.

**Conclusion:** In this real-world, multicenter, retrospective study, the treatment of intracranial aneurysms with the Surpass Evolve FDS was associated with a high rate of complete occlusion and a favorable safety profile. Our findings indicate that complete occlusion was independently predicted by two key factors: periprocedural avoidance of IIb/IIIa inhibitors and a favorable aneurysm morphology without incorporated branches.

## Introduction

Intracranial aneurysms represent a significant cerebrovascular pathology, with a prevalence estimated between 3-5% in the general adult population^1^. While small, unruptured aneurysms may be managed conservatively, larger, symptomatic, or ruptured aneurysms often require intervention to prevent devastating consequences such as subarachnoid hemorrhage^2^. The evolution of endovascular techniques has revolutionized aneurysm treatment^3-5^, with flow-diverting stents (FDs) emerging as a pivotal tool for managing complex, wide-necked saccular aneurysms, or fusiform aneurysms not amenable to traditional coiling or clipping^6-8^.

The Surpass Evolve Flow Diverter System (Stryker Neurovascular, CA, United States) represents a newer generation device designed to promote aneurysm occlusion by reconstructing the parent vessel and inducing intra-aneurysmal thrombosis while maintaining patency of side branches^9,10^. While prior studies, including the SEASE (Surpass Evolve Aneurysm Study) registry^11,12^, have reported on its initial performance, the continuous evaluation through real-world, multicenter experiences is crucial to validate its long-term safety and efficacy across diverse patient populations and clinical practice settings. Such data are essential for refining patient selection, optimizing procedural techniques, and improving clinical outcomes^13^.

This study aims to provide a comprehensive retrospective analysis of the clinical and angiographic outcomes following treatment of intracranial aneurysms with the Surpass Evolve stent across three high-volume leading neurovascular surgical centers in China. We seek to assess its performance in a real-world cohort, identify factors associated with successful occlusion, and contribute to the growing body of evidence guiding its clinical application.

## Methods

### Study Design and Patient Population

This multicenter, retrospective, observational cohort study was reviewed and approved by the Institutional Review Board (IRB) of the coordinating center, with an approval number of 2024235-001. The trial was registered with ClinicalTrials.gov (Identifier: NCT06731946) on 29 November 2024. The requirement for informed consent was waived due to the retrospective design.

Consecutive patients who underwent endovascular treatment of intracranial aneurysms with the Surpass Evolve stent between March 1, 2023, and May 31, 2024, at three tertiary neurovascular treatment centers were identified from prospectively maintained databases.

Inclusion criteria were: (1) age 18 years or older; (2) diagnosis of a saccular, fusiform, blister, or dissecting intracranial aneurysm (ruptured or unruptured); (3) treatment primarily with the Surpass Evolve stent (with or without adjunctive coiling); and (4) availability of at least one follow-up angiogram (Digital Subtraction Angiography [DSA], Magnetic Resonance Angiography [MRA], or Computed Tomography Angiography [CTA]). Exclusion criteria included: (1) treatment for pathologies other than intracranial aneurysms (e.g., arteriovenous malformations); (2) use of other flow-diverting stents as the primary device; or (3) incomplete baseline or follow-up clinical/radiological data.

### Data Collection and Outcome Measures

Demographic, clinical, and procedural data were collected. The primary effectiveness outcome was complete aneurysm occlusion (Raymond-Roy class 1) on the first follow-up angiogram. The primary safety outcome was a composite of major stroke or procedure-related mortality within 30 days. Secondary outcomes included: functional outcome (mRS 0–2), complete occlusion on DSA (O’Kelly-Marotta grade D)^14^, access site complications, minor stroke, fish-mouthing, retreatment, and parent vessel stenosis.

### Imaging and Follow-up Protocol

Angiographic results were assessed by two experienced neuro-interventionalists at each center, with discrepancies resolved by consensus. Follow-up imaging was typically scheduled at 3-6 months and 12-18 months post-procedure, though intervals varied based on clinical practice and aneurysm complexity. The choice of imaging modality (DSA, MRA, or CTA) was at the discretion of the treating team.

### Statistical Analysis

Statistical analysis was performed using SPSS (version 26.0). Continuous variables are presented as median (IQR) and were compared using the Mann-Whitney U test. Categorical variables were compared using the Chi-square or Fisher’s exact test. A two-sided P-value <0.05 was considered significant. Multivariate logistic regression was performed to identify independent predictors of complete occlusion. Variables with P<0.10 in univariate analysis or those deemed clinically relevant were included. Results are reported as adjusted odds ratios (aOR) with 95% confidence intervals (CI). All data generated and analyzed during this study are included in this article.

## Results

### Baseline Patient and Aneurysm Characteristics

A total of 132 patients with 132 target aneurysms were included in the analysis. The median age was 60.0 years (IQR 52.5–69.0), and 80 patients (66.6%) were female. Most patients (98.5%) presented with a favorable baseline functional status (mRS 0–2). Acutely ruptured aneurysms accounted for only 2.3% (n=3) of cases. Common comorbidities included hypertension (48.5%) and hyperlipidemia (37.9%). A comparison with the SEASE study is provided in **Table 1**.

**Table 1.** Patient and aneurysm characteristics.

| Characteristics | SEASE Study | Present Study | P Value |
|---|---|---|---|
| **Patients** | n=305 | n=132 |  |
| Age in years median (IQR) | 59.0 (50.0, 67.0) | 60.0 (52.5, 69.0) | 0.094 |
| Female | 256 (83.9%) | 80 (66.6%) | **<0.001** |
| Baseline modified Rankin scale |  |  |  |
| 0–2 | 291 (96.7%) | 130 (98.5%) | 0.520 |
| 3–4 | 10 (3.3%) | 2 (1.5%) | 0.520 |
| Acutely ruptured | 20 (6.6%) | 3 (2.3%) | 0.080 |
| Comorbidities |  |  |  |
| Hypertension | 149 (49.0%) | 64 (48.5%) | 0.927 |
| Hyperlipidemia | 97 (31.9%) | 50 (37.9%) | 0.218 |
| Diabetes | 33 (10.9%) | 14 (10.6%) | 0.931 |
| Coronary arterial disease | 19 (6.3%) | 2 (1.5%) | **0.030** |
| Stoke or transient ischemic attack | 40 (13.2%) | 31 (23.5%) | **0.007** |
| Smoking (current or past) | 109 (35.9%) | 23 (17.4%) | **<0.001** |
| **Aneurysms** | n=332 | n=132 |  |
| Location |  |  |  |
| Anterior circulation | 316 (95.2%) | 122 (92.4%) | 0.262 |
| Posterior circulation | 16 (4.8%) | 10 (7.6%) | 0.262 |
| ICA | 303 (91.3%) | 103 (78.0%) | **<0.001** |
| ACA | 9 (2.7%) | 8 (6.1%) | 0.085 |
| MCA | 4 (1.2%) | 11 (8.3%) | **<0.001** |
| VA | 13 (3.9%) | 8 (6.1%) | 0.325 |
| BA | 2 (0.6%) | 1 (0.8%) | 1.000 |
| PCA | 1 (0.3%) | 1 (0.8%) | 0.478 |
| Morphology |  |  |  |
| Saccular | 309 (93.1%) | 114 (86.4%) | **0.017** |
| Fusiform | 10 (3.0%) | 11 (8.3%) | **0.010** |
| Blister | 7 (2.1%) | 3 (2.3%) | 1.000 |
| Dissecting | 5 (1.5%) | 4 (3.0%) | 0.272 |
| Pseudoaneurysm | 1 (0.3%) | 0 (0%) | 1.000 |
| Size categories |  |  |  |
| Small (<5 mm) | 161 (48.6%) | 64 (48.5%) | 0.982 |
| Medium (5–10 mm) | 109 (32.9%) | 55 (41.7%) | 0.061 |
| Large (11–25 mm) | 54 (16.3%) | 11 (8.3%) | **0.018** |
| Giant (>25 mm) | 7 (2.1%) | 2 (1.5%) | 1.000 |
| Branch arising from aneurysm | 81 (24.4%) | 27 (20.5%) | 0.176 |

IQR, interquartile range; ICA, internal carotid artery; ACA, anterior cerebral artery; MCA, middle cerebral artery; VA, vertebral artery; BA, basilar artery; PCA, posterior cerebral artery.

Regarding aneurysm characteristics, 122 (92.4%) were located in the anterior circulation, most commonly the internal carotid artery (78.0%). Morphologically, saccular aneurysms predominated (86.4%), followed by fusiform (8.3%). Size distribution: <5 mm (48.5%), 5–10 mm (41.7%), 11–25 mm (8.3%), and >25 mm (1.5%). Significant differences from SEASE were noted in ICA and MCA locations, as well as saccular and fusiform morphologies (**Table 1**).

### Procedural Characteristics

Procedural details are summarized in **Table 2**. Radial access was used in 42.4% of cases (vs. 21.6% in SEASE, P<0.001). A single Evolve stent was implanted in all cases. Balloon angioplasty was used less frequently (7.6% vs. 38.7%, P<0.001). Adjunctive coiling was employed in 9.1% of cases. Intraprocedural use of IIb/IIIa inhibitors was more common (82.6% vs. 9.2%, P<0.001). The most common post-procedural regimen was aspirin plus ticagrelor (56.8%).

**Table 2.** Procedural characteristics.

| **Characteristics** | **SEASE Study** | **Present Study** | **P Value** |
|---|---|---|---|
| Access |  |  |  |
| Femoral | 239 (78.4%) | 76 (57.6%) | **<0.001** |
| Radial | 66 (21.6%) | 56 (42.4%) | **<0.001** |
| Cases with>1 SE implanted | 23 (7.5%) | 0 (0%) | **<0.001** |
| Balloon angioplasty | 118 (38.7%) | 10 (7.6%) | **<0.001** |
| Adjunctive treatments |  |  |  |
| Coils | 20 (6.6%) | 12 (9.1%) | 0.341 |
| Stent | 7 (2.3%) | 10 (7.6%) | **0.008** |
| Intraprocedural IIb/IIIa inhibitor use | 28 (9.2%) | 109 (82.6%) | **<0.001** |
| Post-procedural antiplatelet regimens |  |  |  |
| Aspirin+clopidogrel | 166 (54.6%) | 50 (37.9%) | **0.001** |
| Aspirin+prasugrel | 52 (17.1%) | 0 (0%) | **<0.001** |
| Aspirin+ticagrelor | 85 (28.0%) | 75 (56.8%) | **<0.001** |
| Other | 1 (0.3%) | 7 (5.3%) | **<0.001** |

IQR, interquartile range; SE, Surpass Evolve.

### Safety and Effectiveness Outcomes

The median time to first imaging follow-up was 9.2 months (IQR 5.6–12.4). Primary and secondary outcomes are detailed in **Table 3**.

**Table 3.** Safety and effectiveness outcomes.

| **Characteristics Values** | **SEASE Study** | **Present Study** | **P Value** |
|---|---|---|---|
| **Primary outcomes** |  |  |  |
| Effectiveness |  |  |  |
| Complete aneurysm occlusion (RR 1) | 233/319 (73.0%) | 88/120 (73.3%) | 0.951 |
| Safety |  |  |  |
| Major stroke (ischemic/hemorrhagic) | 6/293 (2.1%) | 3/129 (2.3%) | 0.750 |
| Mortality related to procedure | 2/293 (0.7%) | 0/129 (0%) | 0.500 |
| **Secondary outcomes** |  |  |  |
| Effectiveness |  |  |  |
| Follow-up modified Rankin scale score |  |  |  |
| 0–2 | 282/293 (94.9%) | 125/129 (96.9%) | 0.740 |
| 3–6 | 15/293 (5.1%) | 4/129 (3.1%) | 0.740 |
| Complete aneurysm occlusion on DSA (OKM D) | 166/231 (71.8%) | 82/114 (71.9%) | 0.990 |
| Safety |  |  |  |
| Access site complications | 2/286 (0.7%) | 0/132 (0%) | 0.500 |
| Minor stroke | 9/290 (3.1%) | 8/129 (6.2%) | 0.140 |
| **Additional outcomes** |  |  |  |
| Fish-mouthing |  |  |  |
| Intraoperative | 29/305 (9.5%) | 10/132 (7.5%) | 0.520 |
| At follow-up | 7/211 (3.3%) | 0/120 (0%) | 0.100 |
| Retreatment | 8/286 (2.8%) | 1/120 (0.8%) | 0.450 |
| Parent vessel stenosis at follow-up |  |  |  |
| No stenosis | 153/211 (72.5%) | 111/120 (92.5%) | **<0.001** |
| < 70% stenosis | 53/211 (25.1%) | 4/120 (3.3%) | **<0.001** |
| ≥ 70% stenosis | 3/211 (1.4%) | 4120 (3.3%) | **0.040** |
| Complete occlusion | 2/211 (0.9%) | 1/120 (0.8%) | 1.000 |
| Follow-up characteristics |  |  |  |
| Time to follow-up in months, median (IQR) | 10.2 (6.4, 12.9) | 9.2 (5.6, 12.4) | 0.500 |
| Type of imaging modality |  |  |  |
| Digital subtraction angiography | 211/280 (75.4%) | 114/120 (95.0%) | **<0.001** |
| Magnetic resonance angiography | 40/280 (14.3%) | 0/120 (0%) | **<0.001** |
| CT angiography | 29/280 (10.4%) | 6/120 (5.0%) | 0.060 |

CT, computed tomography; DSA, digital subtraction angiography; IQR, interquartile range; OKM, O’Kelly Marotta scale; RR, Raymond-Roy.

#### Primary Outcomes

##### Effectiveness

The rate of complete aneurysm occlusion (Raymond-Roy class 1) was 73.3% (88/120), which was nearly identical to the SEASE study rate of 73.0% (P=0.951).

##### Safety

Major stroke occurred in 2.3% (3/129) of patients. There were no procedure-related deaths (0% vs. 0.7% in SEASE, P=0.500).

#### Secondary Outcomes

##### Functional Outcome

At the last clinical follow-up, 96.9% (125/129) of patients had a favorable functional outcome (mRS 0–2).

##### Angiographic Outcome (DSA)

On DSA follow-up (available for 114 patients), the rate of complete occlusion (OKM D) was 71.9% (82/114).

##### Complications

Access site complications were rare (0%). The rate of minor stroke was 6.2% (8/129).

##### Stent Apposition and Retreatment

Intraoperative “fish-mouthing” was observed in 7.5% of cases. The retreatment rate was low at 0.8%.

##### Parent Vessel Patency

Follow-up imaging showed no significant parent vessel stenosis in 92.5% of cases. The rate of ≥70% stenosis was 3.3%.

##### Follow-up Imaging Modality

DSA, the predominant follow-up modality (95.0%), was used more frequently than in the SEASE study (75.4%, P<0.001).

##### Case-Control Analysis

Predictors of Complete Occlusion

Patients were categorized into Complete Occlusion (n=88) and Non-Complete Occlusion (n=32). Univariate comparisons are presented in **Table 4**. Intraprocedural use of IIb/IIIa inhibitors was significantly more frequent in the non-occlusion group (93.8% vs. 77.3%, P=0.039). Multivariate analysis showed that non-use of intraprocedural IIb/IIIa inhibitor (aOR 0.38; 95% CI 0.15–0.92; P=0.032) and absence of a branch arising from the aneurysm (aOR 3.57; 95% CI 1.47–8.66; P=0.005) were independent predictors of complete occlusion.

**Table 4.** Case-control study.

| Characteristics | Complete occlusion<br>（N=88） | Non-complete occlusion<br>（N=32） | P Value |
|---|---|---|---|
| **Patients** |  |  |  |
| Age in years median (IQR) | 60.0 (53.0, 67.5) | 61.5 (53.5, 67.0) |  |
| Female | 57 (64.8%) | 21 (65.6%) | 0.931 |
| Baseline modified Rankin scale |  |  |  |
| 0–2 | 86 (97.7%) | 32 (100%) | 1.000 |
| 3–4 | 2 (%) | 0 (0%) |  |
| Acutely ruptured | 2 (2.3%) | 0 (0%) | 1.000 |
| Comorbidities |  |  |  |
| Hypertension | 41 (46.6%) | 18 (56.2%) | 0.347 |
| Hyperlipidemia | 36 (40.9%) | 10 (31.2%) | 0.336 |
| Diabetes | 11 (12.5%) | 3 (9.4%) | 0.637 |
| Coronary arterial disease | 2 (2.3%) | 0 (0%) | 1.000 |
| Stroke or transient ischemic attack | 22 (25.0%) | 6 (18.8%) | 0.473 |
| Smoking (current or past) | 16 (18.2%) | 5 (15.6%) | 0.744 |
| Drinking | 8 (9.1%) | 4 (12.5%) | 0.581 |
| **Aneurysms** |  |  |  |
| Location |  |  |  |
| Anterior circulation | 81 (92.0%) | 30 (93.8%) | 0.754 |
| Posterior circulation | 7 (8.0%) | 2 (6.2%) | 0.754 |
| ICA | 66 (75.0%) | 26 (81.2%) | 0.474 |
| ACA | 7 (8.0%) | 1 (3.1%) | 0.682 |
| MCA | 8 (9.1%) | 3 (9.4%) | 1.000 |
| VA | 5 (5.7%) | 2 (6.2%) | 1.000 |
| BA | 1 (1.1%) | 0 (0%) | 1.000 |
| PCA | 1 (1.1%) | 0 (0%) | 1.000 |
| Morphology |  |  |  |
| Saccular | 78 (88.6%) | 26 (81.2%) | 0.292 |
| Fusiform | 6 (6.8%) | 5 (15.6%) | 0.140* |
| Blister | 2 (2.3%) | 0 (0%) | 1.000 |
| Dissecting | 2 (2.3%) | 1 (3.1%) | 0.550 |
| Size categories |  |  | 0.384 |
| Small (<5 mm) | 44 (50.0%) | 18 (56.2%) |  |
| Medium (5–10 mm) | 37 (42.0%) | 10 (31.2%) |  |
| Large (11–25 mm) | 5 (5.7%) | 4 (12.5%) |  |
| Giant (>25 mm) | 2 (2.3%) | 0 (0%) |  |
| Aneurysmal neck ≥4mm | 43 (48.9%) | 15 (46.9%) | 0.847 |
| Branch arising from aneurysm | 12 (13.6%) | 12 (37.5%) | **0.004\*** |
| **Procedure** |  |  |  |
| Access |  |  |  |
| Femoral | 50 (56.8%) | 17 (53.1%) | 0.719 |
| Radial | 38 (43.2%) | 15 (46.9%) | 0.719 |
| Balloon angioplasty | 9 (10.2%) | 0 (0%) | 0.062* |
| Adjunctive treatments |  |  |  |
| Coils | 10 (11.4%) | 1 (3.1%) | 0.283 |
| Stent | 8 (9.1%) | 1 (3.1%) | 0.442 |
| Intraprocedural IIb/IIIa inhibitor use | 68 (77.3%) | 30 (93.8%) | **0.039\*** |
| Post-procedural antiplatelet regimens |  |  | 0.275 |
| Aspirin+clopidogrel | 34 (38.6%) | 8 (25.0%) |  |
| Aspirin+ticagrelor | 49 (55.7%) | 23 (71.9%) |  |
| Other | 5 (5.7%) | 1 (3.1%) |  |

\* Variables with P < 0.10 in univariate analysis (marked with an asterisk) were included in a multivariate logistic regression model to identify independent predictors of complete aneurysm occlusion. The final model was constructed using a backward stepwise selection method and adjusted for covariates including Fusiform morphology, Branch arising from aneurysm, Balloon angioplasty, and Intraprocedural IIb/IIIa inhibitor use.
Multivariate analysis results: After adjustment, non-use of an intraprocedural IIb/IIIa inhibitor (adjusted odds ratio [aOR] 0.38; 95% confidence interval [CI] 0.15–0.92; P=0.032) and absence of a branch arising from the aneurysm (aOR 3.57; 95% CI 1.47–8.66; P=0.005) remained independent predictors of complete occlusion. Fusiform morphology was not significantly associated with the occlusion outcome in the final model (aOR 0.48; 95% CI 0.17–1.34; P=0.162). The variable Balloon angioplasty was excluded from the final model due to a lack of events (0%) in the non-complete occlusion group, which prevented stable model estimation. The overall model fit was adequate (Hosmer-Lemeshow test P=0.42; Nagelkerke R² = 0.18).

## Discussion

This multicenter retrospective study provides a real-world assessment of the Surpass Evolve Flow Diverter System for the treatment of intracranial aneurysms in the Chinese population. The key findings are: (1) a high and reproducible complete occlusion rate of 73.3%, consistent with prior registry data; (2) a favorable safety profile with low rates of major stroke (2.3%) and no procedure-related mortality; (3) excellent functional outcomes, with 96.9% of patients achieving an mRS of 0–2 at follow-up; and (4) the identification of both procedural (non-use of IIb/IIIa inhibitor) and morphological (absence of a branch arising from the aneurysm) factors as independent predictors of complete occlusion.

The observed complete occlusion rate aligns remarkably well with the 73.0% reported rate in the SEASE study^11^, suggesting robust and consistent performance of the Surpass Evolve stent across different study populations and practice settings. This rate is also comparable to occlusion rates reported for other established flow-diverting devices reinforcing the class effect of flow diversion^15^,. The high rate of favorable functional outcomes further supports the clinical efficacy of this treatment strategy, indicating that successful aneurysm occlusion is associated with preserved or improved neurological function in the vast majority of patients^16^..

The safety profile demonstrated in this cohort is encouraging. The absence of procedure-related mortality and the low major stroke rate are critical indicators for the risk-benefit assessment of flow diversion^17^. The higher utilization of radial access in our cohort (42.4%) reflects a modern shift in neurointerventional practice and may contribute to reduced access site complications^18^. The significant reduction in the use of balloon angioplasty and multiple stents compared to the SEASE study may reflect increasing clinicians’ experience and confidence in the device’s deliverability and opening characteristics, potentially contributing to improved safety^11^.

The analysis of parent vessel stenosis at follow-up revealed excellent patency, with 92.5% of cases showing no stenosis. The rate of ≥70% stenosis was low (3.3%). These results are favorable and suggest that the Evolve stent has a good biocompatibility profile, with a low incidence of significant intimal hyperplasia leading to stenosis in the short-to-medium term^19^.

Our analysis identified two independent predictors of treatment success. First, the independent association between non-use of an intraprocedural IIb/IIIa inhibitor and complete aneurysm occlusion requires careful interpretation. While these agents are commonly used to manage acute intraprocedural thrombosis^20,21^, their potential impact on long-term occlusion is complex^22,23^. The association between IIb/IIIa inhibitor use and a lower rate of complete occlusion in our analysis requires careful interpretation. It may reflect a clinical scenario where these agents were selectively administered in more complex or high-risk procedures perceived to have a greater immediate thromboembolic risk (e.g., those with visible thrombus, significant stent manipulation, or in ruptured cases), which themselves are associated with poorer long-term occlusion outcomes^24^. Although our multivariate analysis attempted to adjust for several confounding variables, residual bias or unmeasured confounding factors (such as specific procedural complexities^25^ or thrombus burden^26^) cannot be excluded. This finding underscores the complexity of periprocedural antiplatelet management in flow diversion^14^. It suggests that routine or liberal use of potent intravenous antiplatelets during the procedure may not be universally beneficial for achieving long-term occlusion^27^, and a more tailored, risk-stratified approach warrants investigation in future studies^28^.

Second, the absence of a branch arising from the aneurysm was a strong independent predictor of complete occlusion. This aligns with the mechanistic principle of flow diversion, which relies on flow disruption and stasis within the aneurysm sac to induce thrombosis^6,19^. Computational and clinical studies have shown that the presence of a patent branch vessel originating from the aneurysm dome or neck can provide persistent inflow or outflow, sustaining intra-aneurysmal flow and thereby hindering complete thrombosis and healing^29,30^. This morphological feature has been consistently suggested as a potential challenge for flow diverter treatment, associated with lower occlusion rates and higher retreatment needs^31,32^. Our results reinforce the importance of careful aneurysm morphology assessment during treatment planning, as aneurysms with incorporated branches might require longer follow-up.

## Limitations

This study has several limitations inherent to its retrospective, non-randomized design. First, the lack of a control group treated with alternative devices or strategies limits comparative effectiveness conclusions. Second, treatment decisions, including antiplatelet regimens and the use of IIb/IIIa inhibitors, were not standardized but followed institutional protocols, introducing potential confounding. Third, the median follow-up duration of approximately 9 months, while informative, is relatively short for assessing the long-term durability of occlusion and late complications. Finally, despite being multicenter, the sample size remains moderate, and the study may be underpowered to detect rare adverse events or subtle predictors of outcome.

## Conclusion

In this multicenter, real-world experience, endovascular treatment of intracranial aneurysms with the Surpass Evolve Flow Diverter System was associated with a high rate of complete occlusion, a favorable safety profile, and excellent functional outcomes. Successful occlusion was independently associated with the non-use of intraprocedural IIb/IIIa inhibitors and the absence of a branch arising from the aneurysm. These observations highlight the interplay between procedural medication management and aneurysm architecture in determining treatment outcome, and merit further prospective evaluation to optimize therapeutic strategies.

## Data Availability

All data generated and analyzed during this study are included in this article.

